# A multisite genomic epidemiology study of *Clostridioides difficile* infections in the U.S. supports differential roles of healthcare versus community spread for two common strains

**DOI:** 10.1101/2020.11.28.20240192

**Authors:** Arianna Miles-Jay, Vincent B. Young, Eric G. Pamer, Tor C. Savidge, Mini Kamboj, Kevin W. Garey, Evan S. Snitkin

**Author notes:** **Corresponding author:** Evan S. Snitkin, 1520D MSRB I, 1150 W. Medical Center Dr., Ann Arbor, MI, 48109. **Repositories:** All whole genome sequence data was uploaded to the National Center for Biotechnology Information (NCBI) Sequence Read Archive (SRA) under Bioproject accessions PRJNA595724, PRJNA561087, and PRJNA594943.

## Abstract

*Clostridioides difficile* is the leading cause of healthcare-associated infectious diarrhea. However, it is increasingly appreciated that healthcare-associated infections derive from both community and healthcare transmission, and that the primary sites of *C. difficile transmission* may be strain dependent. We conducted a multisite genomic epidemiology study to assess differential genomic evidence of healthcare vs. community spread for two of the most common *C. difficile* strains in the U.S.: sequence type (ST) 1 (associated with Ribotype 027) and ST2 (associated with Ribotype 014/020). Isolates recovered from stool specimens collected during standard clinical care at three geographically distinct U.S. medical centers between 2010 and 2018 underwent whole genome sequencing and phylogenetic analyses. ST1 and ST2 isolates both displayed some evidence of phylogenetic clustering by study site, but clustering was stronger and more apparent in ST1, consistent with our healthcare-based study more comprehensively sampling local transmission of ST1 compared to ST2 strains. Analyses of pairwise single nucleotide variant (SNV) distance distributions were also consistent with more evidence of healthcare transmission of ST1 compared to ST2, with 44% of ST1 isolates being within 2 SNVs of another isolate from the same geographic collection site compared to 5.5% of ST2 isolates (p-value = <0.001). Conversely, ST2 isolates were more likely to have close genetic neighbors across disparate geographic sites compared to ST1 isolates, further supporting non-healthcare routes of spread for ST2 and highlighting the potential for misattributing genomic similarity among ST2 isolates to recent healthcare transmission. Finally, we estimated a lower evolutionary rate for the ST2 lineage compared to the ST1 lineage using Bayesian timed phylogenomic analyses, and hypothesize that this may contribute to observed differences in geographic concordance among closely related isolates. Together, these findings suggest that ST1 and ST2, while both common causes of *C. difficile* infection in hospitals, show differential reliance on community and hospital spread. This conclusion supports the need for strain-specific criteria for interpreting genomic linkages and emphasizes the importance of considering differences in the epidemiology of circulating strains when devising interventions to reduce the burden of *C. difficile* infections.

**DATA SUMMARY:** All whole genome sequence data was uploaded to the National Center for Biotechnology Information (NCBI) Sequence Read Archive (SRA) under Bioproject accessions PRJNA595724, PRJNA561087, and PRJNA594943. Metadata that comply with patient privacy rules are included in the Supplementary Materials.

## INTRODUCTION

*Clostridioides difficile* is a gram-positive spore-forming anaerobic bacterium that is a dominant cause of infectious diarrhea, colitis, and colitis-associated death in the United States [1,2]. While *C. difficile* infection (CDI) is classically considered nosocomial [3], recent molecular epidemiologic research suggests that less than 40% of CDI cases are linkable to other symptomatic CDI cases within the same hospital [4–6]. This insight has disrupted the paradigm of *C. difficile* as an exclusively nosocomial pathogen and expanded interest into the roles of alternative routes of *C. difficile* transmission, including community-based acquisition with subsequent progression to CDI within healthcare settings [7].

Different *C. difficile* strains may have varying propensities for transmission within healthcare vs. the community, and fluroquinolone resistance has been raised as a potential defining characteristic of strains that spread more readily within healthcare settings [8]. In particular, the largely fluoroquinolone-resistant (FQR) Ribotype (RT) 027—also known as NAP1 via pulse-field gel electrophoreses or sequence type (ST) 1 via multi-locus sequence typing (MLST)—has been implicated in numerous hospital-based CDI outbreaks and is most commonly healthcare-associated according to surveillance definitions based on past hospitalizations [9–12]. Another common *C. difficile* lineage in the U.S., RT014/020 (corresponding to STs 2, 49, and 13), is largely fluoroquinolone sensitive (FQS) and, while it is frequently characterized as healthcare-associated using these same surveillance definitions, has not been associated with hospital-based outbreaks [13]. Associations between *C. difficile* strain type and propensity for healthcare-associated transmission would indicate that devising effective interventions for reducing the burden of CDI may require an understanding of the molecular epidemiology of locally circulating strains, and that strain-specific incidence may be a more accurate metric of the successful prevention of *C. difficile* transmission within hospitals.

Whole genome sequencing (WGS) can provide insight into the potential contribution of healthcare vs. community spread of particular strains, even in the absence of comprehensive sampling of transmission networks. Recent studies that applied WGS to European clinical *C. difficile* isolates found that RT027/ST1 displayed genomic patterns consistent with healthcare-associated-spread, while RT014/020/ST2 displayed genomic patterns more consistent with community-associated reservoirs [6,8]. However, these distinct epidemiologic patterns have not yet been assessed using genomic data gathered from U.S.-based *C. difficile* isolates. Here, we applied WGS to isolates collected from three geographically distinct U.S. medical centers to assess differential genomic evidence of healthcare vs. community spread between two of the most common *C. difficile* strains: ST1 and ST2.

## METHODS

### Data collection

New *C. difficile* sequences were derived from clinical stool specimens collected as part existing molecular surveillance programs that took place at three U.S. medical centers: Michigan Medicine (UM) between 2010 and 2013 [14], Texas Medical Center Hospitals (TMC) between 2011 and 2017 [15], and Memorial Sloan Kettering Cancer Center (MSKCC) between 2013 and 2017 [16]. At all three sites, toxigenic *C. difficile* positive stool specimens were collected, *C. difficile* isolates were recovered from the speciemens, and DNA was extracted from a single colony as previous described [14–16]. Isolates underwent molecular typing via ribotyping at UM and TMC [17], and MLST at MSKCC [18]. DNA from a sample of isolates that were typed as RT027 or RT014/020 at UM and TMC and ST1 or ST2 at MSKCC was extracted for whole genome sequencing. The Institutional Review Boards at each of the study sites approved the study protocols.

### Whole genome sequencing and bioinformatic methods

DNA was sent to UM and the Nextera XT library preparation kit (Illumina, San Diego, CA) was used to prepare sequencing libraries according to the manufacturer’s instructions. WGS was executed on an Illumina Hiseq platform with 150 base-pair paired-end reads and a targeted read depth of >100X. Sequence data are available from the National Center for Biotechnology Information (NCBI) Sequence Read Archive (SRA) under BioProjects PRJNA595724, PRJNA561087, and PRJNA594943. The bioinformatics methods applied to the new *C. difficile* sequences to identify single nucleotide variants (SNVs) and build phylogenetic trees were executed as previously described [19]. Briefly, raw sequencing reads were trimmed using Trimmomatic to remove low quality bases and adapter sequences. Trimmed reads were then mapped to existing complete reference genomes within the same ST (R20291 for ST1 [GenBank accession number FN545816], and W0022a for ST2 [GenBank accession number CP025046]) with the Burrows-Wheeler short read aligner [20–22]. PCR duplicates were discarded and variants were called using SAMtools mpileup and bcftools [23]. Gubbins was used to remove variant sites located in putative recombinant regions [24]. *In silico* multilocus sequence typing (MLST) was performed using ARIBA and only isolates that were identified as ST1 and ST2 were included in all analyses [25]. Maximum-likelihood phylogenies were built using IQ-TREE with a generalized time reversible nucleotide substitution model; phylogenies were rooted using *C. difficile* 630 as an outgroup (GenBank accession number GCA_000009205.2)[26,27]. Fluroquinolone resistance was assigned based off of the presence of previously identified fluroquinolone resistance-associated gryA and gyrB alleles [28]. ST1 isolates were further classified into previously identified FQS, FQR1, and FQR2 lineages by examining how new isolates clustered with publicly available FQR1 and FQR2 isolates [29].

### Evaluation of phylogenetic clustering

To compare the level of clustering by geographic collection site between newly sequenced ST1 and ST2 isolates, we overlaid geographic collection site onto the maximum-likelihood whole genome phylogenies and applied a previously described approach for formal clustering assessment [30]. First, we tabulated the number of isolates in a “pure” subtree of each phylogeny—defined as a subtree made up of 2 or more isolates collected from a single geographic site that was found in >90% of bootstrapped phylogenies. To determine whether this number was different than would be observed by chance given the phylogenetic topology and location frequency, we calculated an empirical p-value by randomizing geographic labels and re-calculating this number 1000 times.

### Evaluation of evidence of recent transmission

Evidence of recent transmission was assessed using pairwise SNV distance matrices and two analytic approaches. First, we compared the lower tail (5^th^ percentile) of the distribution of pairwise SNV distances of pairs of isolates collected from the same collection site to that same metric among pairs of isolates collected from different collection sites by calculating a 5^th^ percentile SNV-distance ratio (5^th^ percentile SNV distance within sites/5^th^ percentile SNV distance between sites). To assess whether this ratio indicated an enrichment of close linkages within collection sites greater than could be expected by chance, we randomly permuted collection sites and re-calculated the ratio 10,000 times; an observed ratio below the 2.5% percentile of the distribution of expected ratios was applied to support significant enrichment of close genetic linkages within study sites. Second, we classified genomic linkages using an SNV-distance threshold of 2 SNVs and compared the proportion of genomically linked isolates (defined as being linked to at least one other isolate) among ST1 isolates compared to those among ST2 isolates using chi-squared tests. An SNV threshold of 2 SNVs is commonly used to identify pairs of *C. difficile* isolates that were likely related via direct transmission/acquisition from a common source; this threshold is based off of evolutionary rates estimated from within-host evolution [4]. We then assessed the sensitivity of these results to larger thresholds of 5-10 SNVs. We also compared the proportion of isolates genomically linked to at least one isolate collected from a different geographic collection site between ST1 and ST2 using chi-squared tests. All analyses were completed in R v4.0.2.

### Estimation of evolutionary rates

We applied Bayesian timed phylogenomic analyses in order to estimate and compare evolutionary rates between ST1 and ST2 lineages using BEAST v1.10.4 [31]. To increase the power of timed phylogenomic analyses, existing ST1 and ST2 whole genome sequences were downloaded from the NCBI SRA; isolates were selected from a recent publication that compiled isolates from several previous *C. difficile* genome collections along with their ST and sampling date [32]. The combined collection of existing and new sequences was then pared down to facilitate running Bayesian phylogenomic analyses. First, in an effort to maximize genetic diversity, one randomly selected isolate from each pair of isolates within 2 SNVs of one another was removed. Second, isolates from overrepresented geographic locations were randomly downsampled until the total number of isolates was less than 425. The final list of isolates that were included in these analyses can be found in Supplementary Table 1.

We assessed the suitability of the data for timed phylogenomic analyses by examining temporal signal—or the relationship between genomic differences and sampling date—using two methods. First, we examined a regression of sampling time vs. root-to-tip genetic distance using Tempest and BactDating [33,34]. We then formally evaluated temporal signal using date randomization tests, randomly permuting the sampling dates 10 times and comparing the evolutionary rate estimates and their 95% credible intervals for the random datasets to the estimates from the real data. We report both the more relaxed and more strict criteria for temporal signal assessment using this approach: with the more relaxed criteria being met if the estimated evolutionary rate was not included in the 95% credible intervals of 10 date randomized datasets (CR1), and the more strict being met if the 95% credible interval of the estimated evolutionary rate did not overlap any of the 95% credible intervals of the date randomized datasets (CR2) [35]. We proceeded with evolutionary rate estimates so long as the data met CR1.

To select BEAST model assumptions for both the date randomization tests and the final evolutionary rate estimates, we started with a general time reversible nucleotide substitution model with gamma distributed rate heterogeneity and the simplest clock and demographic model assumptions: a strict molecular clock and constant demographic prior. We then systematically examined to what extent the data violated the strict clock and constant demographic model prior assumptions and thus, to what extent more complex models were warranted. To assess whether the data violated a strict clock assumption, we evaluated whether the coefficient of variation parameter in the models with an uncorrelated relaxed lognormal clock had a 95% highest posterior density interval (HPD) that overlapped 0; if not, we used this as evidence of the assumptions of a strict clock being violated and applied an uncorrelated relaxed lognormal clock with a lognormal prior distribution with a mean of 5.0×10^−7^ and standard deviation of 8×10^−7^ based on previous evolutionary rate estimates (while allowing still allowing for significant deviation) [29,36]. To assess to what extent the data violated a constant demographic model, we ran models with exponential growth demographic model prior, and evaluated whether the 95% credible interval of the exponential growth rate parameter overlapped 0. If the exponential growth rate parameter was substantially different from 0, we attempted running a more flexible but parameter rich Gaussian Markov Random Field (GMRF) skyride model, which allows for periods of growth as well as periods of stasis [37]. For each model, a Markov-chain Monte Carlo was run for 200 million generations and sampled every 10,000 iterations; a Tempest-rooted starting tree was included in all runs to accelerate convergence [33]. All ESS values were checked for being above 200 using Tracer after removing the first 10% of steps as burn-in [38].

## RESULTS

There were 382 new whole genome sequences generated from the 3 U.S. study sites located in Michigan, Texas, and New York; 199 ST1 and 183 ST2 (Supplementary Figure 1). The majority of ST1 isolates were FQR, relatively evenly distributed between the previously described FQR1 and FQR2 lineages, and the FQS isolates clustered together in one ancestral clade. Conversely, ST2 isolates were largely FQS, with FQR isolates occurring in a two small clusters as well as singletons scattered throughout the phylogeny (Figure 1). ST1 sequences were less diverse than ST2 sequences: after quality and recombination filtering, the ST1 alignment consisted of 1108 SNVs (median pairwise SNV distance 35, range 0-85), while the ST2 alignment consisted of 2119 SNVs (median pairwise SNV distance 52, range 1-156) (Figure 2).

**Figure 1:**
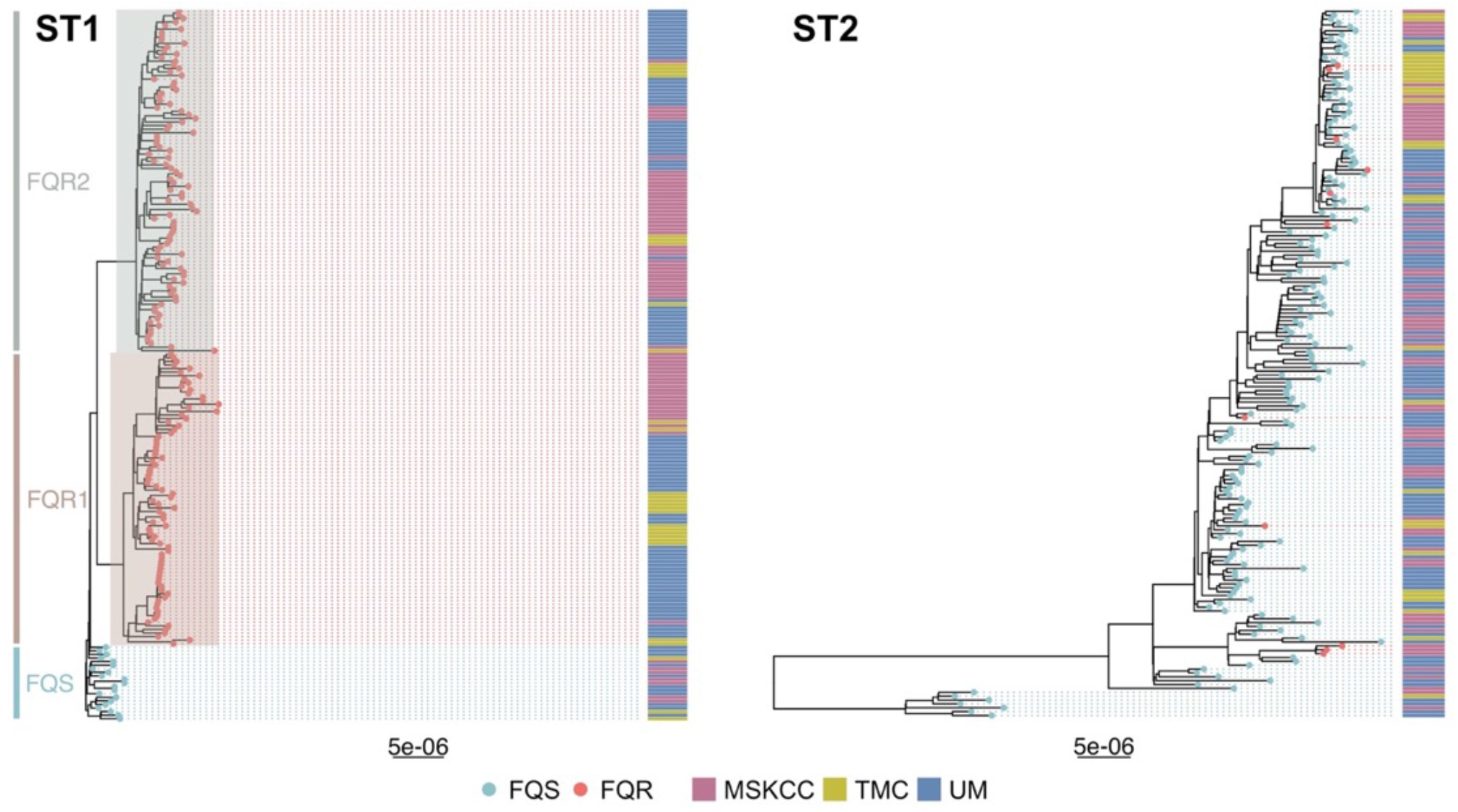
Maximum likelihood phylogenetic trees of newly sequenced *C. difficile* isolates that are ST1 and ST2. Tips are colored by fluroquinolone-resistant (FQR) vs. fluroquinolone-sensitive (FQS) as determined by the presence of previously identified fluroquinolone-resistance-associated gryA and gyrB alleles. Previously identified ST1 lineages (FQS, FQR1, and FQR2) are highlighted, collection site is included in an adjacent heatmap. Tree scales are in single nucleotide changes per quality- and recombination-filtered site.

**Figure 2:**
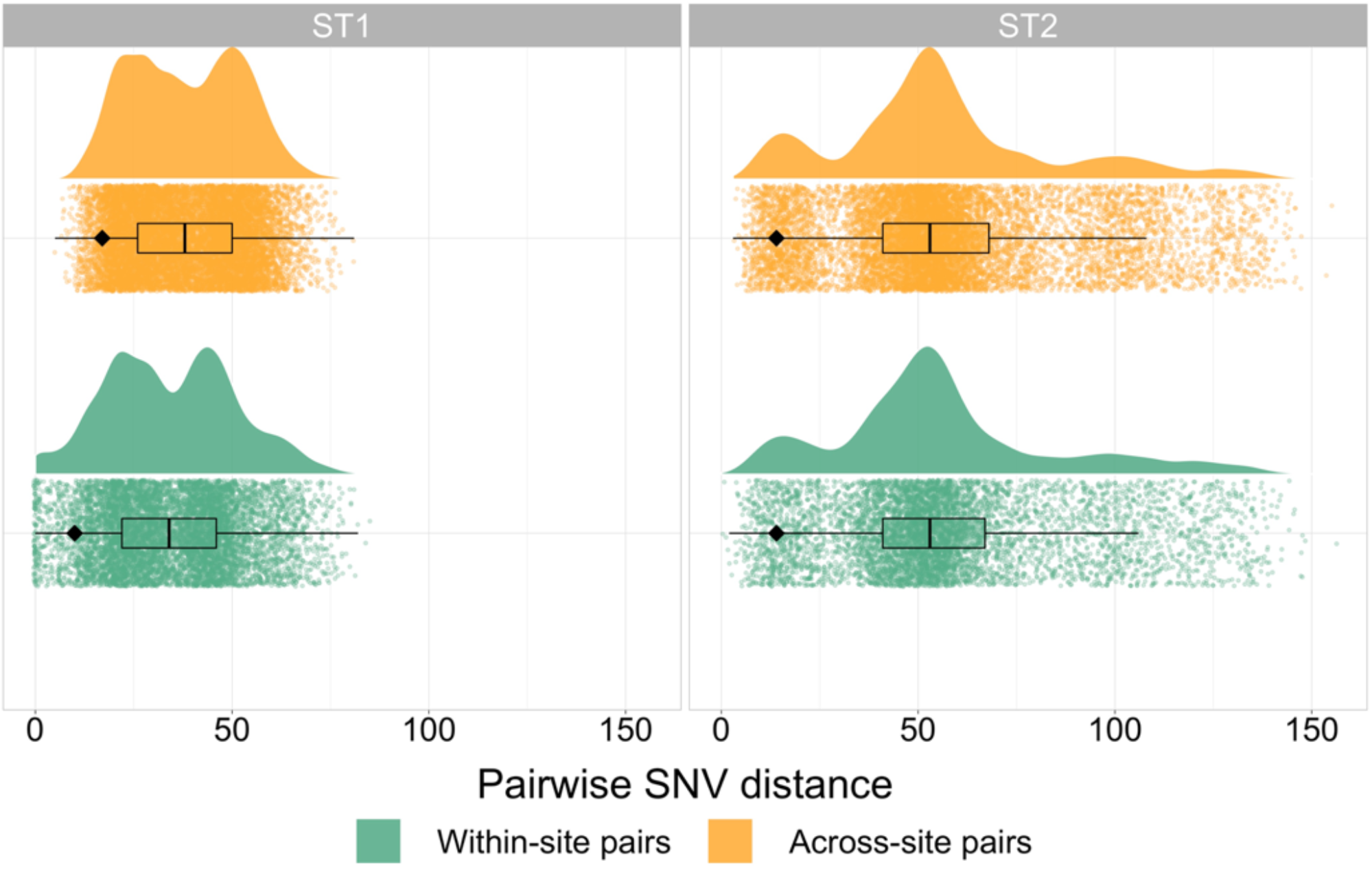
Pairwise single nucleotide variant (SNV) distribution between pairs of isolates from the same collection site vs. pairs of isolates from geographically distinct collection sites for both ST1 and ST2. The black diamond indicates fifth percentile SNV distances for each category.

### ST1 exhibits stronger evidence of phylogenetic clustering by geography compared to ST2

To begin our comparison of ST1 and ST2 isolates, we first examined the association between phylogenetic and geographic structure by overlaying the geographic site each isolate was collected from onto strain-specific whole genome phylogenies. Visual examination of these phylogenies revealed a striking difference in geographic clustering, with ST1 displaying larger clusters and ST2 displaying more numerous, smaller clusters and more geographic mixing (Figure 1). The exception to this observation was the FQS ST1 clade, which appeared more geographically mixed than the FQR ST1 clades. While statistical assessments demonstrated that both ST1 and ST2 displayed more evidence of geographic clustering than would be expected to occur by chance (empiric p-values both <0.001), clustering was more non-random for ST1 than ST2 (Supplementary Figure 2). This enhanced geographic clustering among ST1 isolates could reflect that our healthcare-based study more completely sampled local transmission networks among ST1 isolates compared to ST2 isolates, or it could reflect ST1 spreading via more localized community or healthcare reservoirs with minimal long-distance transmission.

### ST1 isolates display more evidence of recent transmission than ST2, while ST2 isolates are more likely to share intermediate genetic linkages across disparate geographic sites

To further investigate whether plausible healthcare-associated transmission among ST1 isolates was driving the geographic clustering patterns we saw in the phylogenies, we next examined the prevalence and nature of close genetic linkages within each lineage as captured by pairwise SNV distances. Isolates linked by very small SNV distances are plausibly linked via recent transmission, and we would expect our healthcare-based study to more comprehensively sample healthcare-associated transmission than community-associated transmission. When examining the SNV distance distributions between and within collection sites, among ST1 isolates, we observed more closely related pairs of isolates from the same geographic collection site (reflected by a heavier lower tail of the distribution) compared to pairs of isolates collected from different geographic collection sites (5^th^ percentile SNV distance within sites/5^th^ percentile SNV distance between sites = 0.59, expected ratio 95% interval 0.93-1.00, Figure 2). However, we did not observe this same pattern among ST2 isolates (5^th^ percentile SNV distance within sites/5^th^ percentile SNV distance between sites = 1.00, expected ratio 95% interval 0.93-1.00, Figure 2). Application of SNV distance thresholds demonstrated that 88 (44%) ST1 isolates were within 2 SNVs of another isolate from the same geographic collection site compared to 10 (5.5%) ST2 isolates (p-value = <0.001). As the SNV threshold was increased to intermediate values of 5 and 10 SNVs, this trend was maintained (all p < 0.001, Figure 3A). Conversely, at the 5 and 10 SNV thresholds, linked ST2 isolates were more likely to be linked to an isolate from a different geographic collection site compared to linked ST1 isolates (all p < 0.001, Figure 3A). These geographically discordant intermediate genomic linkages among ST2 were not associated with temporal linkages, with the days between sample collection ranging from 6 to 2,479 days (Figure 3B). Among geographically discordant ST1 isolates pairs, FQS isolates were overrepresented, with the only pair of geographically discordant ST1 isolates linked within 5 SNVs being FQS and 14/31 (45.2%) geographically discordant ST1 isolates linked within 10 SNVs being FQS even though FQS isolates made up only 21/199 (10.6%) of isolates overall. Together, these findings are consistent with evidence of recent healthcare transmission among ST1 isolates and transmission outside of the hospital among ST2 isolates, and also raise questions about the underlying reasons why ST2 isolates are more likely to be closely related across disparate geographic sites.

**Figure 3:**
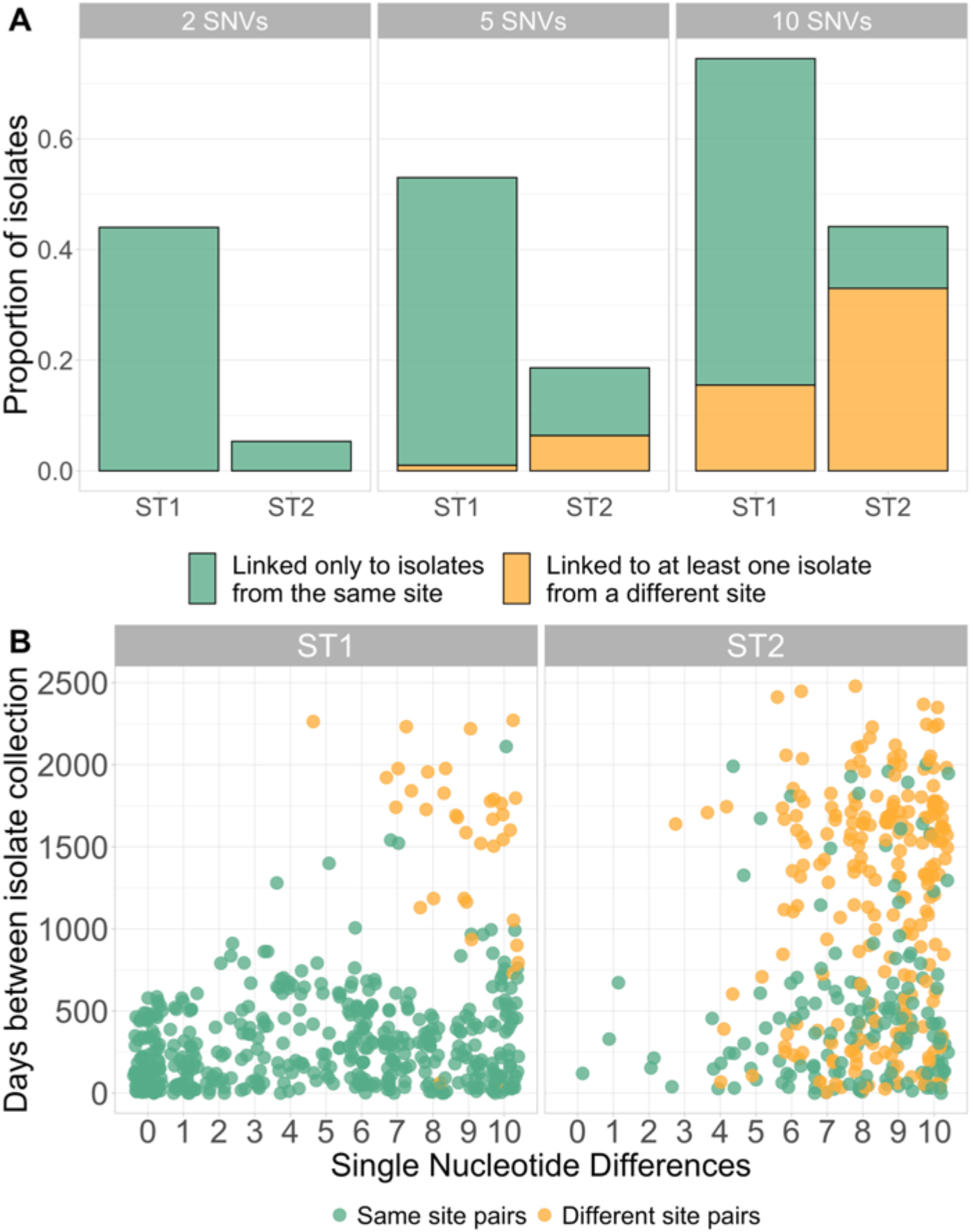
A) Bar plot showing the proportion of ST1 and ST2 isolates that are genomically linked to another isolate, either from the same collection site (green) only or from at least one different collection site (orange), at varying SNV thresholds. B) Scatter plot of days between collection and pairwise SNV distance up to 10 SNVs, where each dot represents one pair of isolates. Points are colored by whether they are collected from the same geographic collection site (green) or different geographic collection sites (orange). Points are jittered to improve clarity.

### Timed phylogenomic analyses demonstrate evidence of evolutionary rate heterogeneity within and between ST1 and ST2 lineages

Our observation that ST2 is more likely to be genomically linked at intermediate SNV thresholds across disparate geographic sites compared to ST1 isolates led us to explore the potential mechanisms underlying this difference. Two factors we hypothesized might contribute to these findings are 1) increased transmission of ST2 via community-based reservoirs that facilitate more rapid spread over large geographic distances and/or 2) a slower average evolutionary rate among ST2 isolates resulting in less genetic changes over larger amounts of time and space. While examining the former hypothesis was beyond the scope of this study, we explored the plausibility of the latter hypothesis by estimating evolutionary rates for ST1 and ST2 using the BEAST Bayesian phylogenetic software [31]. There were 418 ST1 and 418 ST2 isolates included in this analysis; sequences included a mix of newly sequenced and publicly available global genomes in order to maximize temporal and genetic diversity while maintaining a sample size manageable by the BEAST software (Supplementary Table 1). For ST1 isolate selection, we also opted to maintain all FQS ST1 isolates, given our observations that they may display distinct epidemiological patterns from FQR ST1 isolates.

Temporal signal analyses, while initiated as a necessary precursor to timed phylogenomic analyses in BEAST, revealed interesting differences between the clock-like nature of ST1 and ST2 isolates. While root-to-tip regression analyses suggested similarly weak but sufficient temporal signal to proceed with timed phylogenomic analyses in BEAST (indicated by positive correlation coefficients, Supplementary Figure 3), the more rigorous hypothesis testing date randomization tests demonstrated more evidence of temporal signal among ST1 isolates, which passed both the more relaxed CR1 and more stringent CR2 criteria for temporal analyses, compared to ST2 isolates, which passed CR1 but not CR2 (Supplementary Figure 4). The root-to-tip regression also highlighted different temporal patterns among FQS-ST1 isolates compared to FQR-ST1 isolates, which was observed again in date randomization tests on FQS-ST1 and FQR-ST1 isolates separately; the FQR-ST1 isolates appeared to drive the temporal signal in the data, and when considered alone, FQS-isolates were more like ST2 isolates, passing the more relaxed CR1 temporal signal criteria but not the more stringent CR2. This observation was consistent with our pairwise SNV distance findings of distinct patterns among FQS ST1 isolates, and motivated conducting further analyses both with all ST1 isolates together as well as with FQR ST1 isolates (n = 359) and FQS ST1 isolates (n = 59) considered separately.

All datasets demonstrated evidence of evolutionary rate heterogeneity throughout the phylogeny, resulting in the application of uncorrelated relaxed lognormal molecular clock models along with a constant demographic priors (see Supplementary Figures 5-6 and Supplementary Results for details). Overall, when considering all ST1 isolates together compared to all ST2 isolates, evolutionary rate estimates were slightly higher for ST1 compared to ST2, although the 95% credible intervals overlapped. However, ST1’s faster evolutionary rate was driven by FQR ST1 isolates; when separating out FQS and FQR ST1 isolates, the FQR ST1 evolutionary rate estimates emerged as significantly higher than that of ST2 isolates (with non-overlapping 95% credible intervals) while FQS ST1 isolates had similar evolutionary rate estimates to ST2 isolates (Figure 4). These evolutionary rates translate to approximately 1.36 (95% credible interval 1.20-1.52) nucleotide changes per year for FQR ST1, 0.80 (95% credible interval 0.51-1.08) nucleotide changes per year for FQS-ST1, and 0.89 (95% credible interval 0.74-1.05) nucleotide changes per year for ST2. These results are consistent with the hypothesis that a slightly slower average evolutionary rate among ST2 and FQS ST1 isolates compared to FQR ST1 isolates might contribute to our observed discordance between genomic and epidemiologic linkages among those isolates.

**Figure 4:**
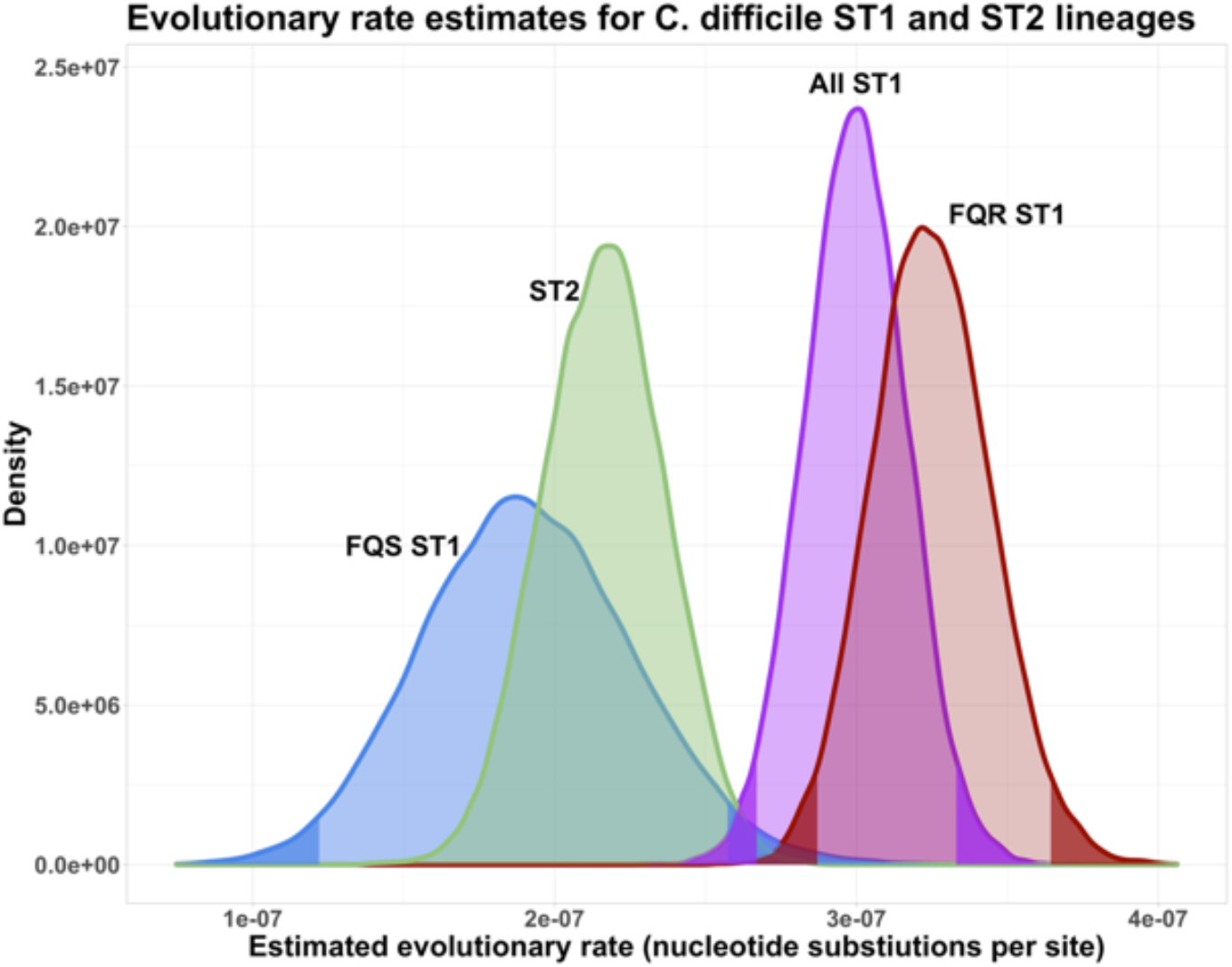
Posterior probability density of the evolutionary rates estimates for *C. difficile* ST1 and ST2 lineages, with ST1 isolates considered together as well as separated out into FQR-ST1 and FQS-ST1 isolates. Dark shaded areas of the density curves indicate the lower 2.5% and upper 97.5% of the distributions; light shaded areas indicate 95% credible intervals. Evolutionary rates are considered significantly different from one another when the 95% credible intervals of their posterior probability densities do not overlap.

## DISCUSSION

In this study, we investigated the genomic epidemiology of two dominant *C. difficile* lineages, ST1 and ST2, across three geographically distinct U.S. medical centers. We observed more genomic evidence of geographic clustering and recent transmission among ST1 isolates compared to ST2 isolates, while also finding more linkages among ST2 isolates from disparate geographic collection sites at intermediate genomic linkage thresholds. Lastly, we estimated a slightly more rapid average evolutionary rate for FQR ST1 isolates compared to FQS ST1 isolates and ST2 isolates using Bayesian timed phylogenomic methods.

Previous studies have reported both more evidence of broad geographic clustering [8] and more evidence of recent transmission within healthcare settings [6] among European ST1 *C. difficile* isolates compared to other types of *C. difficile*. To our knowledge, these are the first U.S.-based multisite data to support these findings. Our observations are consistent with ST1 being associated with hospital outbreaks [9–11], being the most predominant healthcare-associated *C. difficile* strain according to surveillance definitions based on timing since last healthcare exposure [13], and being more prevalent in hospital than community environmental sampling [39]. The factors contributing to increased spread of ST1 within healthcare are not well defined, however, fluroquinolone resistance has been proposed as a driving feature. In support of this, Eyre et al. noted that other FQR *C. difficile* strains were also more likely to cluster by country compared to FQS *C. difficile* strains [13]. Our observations of distinct epidemiological and evolutionary patterns among FQS compared to FQR ST1 isolates are also consistent with this hypothesis. If within-healthcare transmission is the dominant mode of ST1 spread, infection control interventions and antimicrobial-stewardship within healthcare should jointly reduce the incidence of CDI due to ST1. Such reductions have been reported in the UK after implementation of national infection prevention and antimicrobial stewardship policies [40].

Conversely, ST2 seems to have followed a different route to pathogenic success. RT014/ST2 has been reported as one of the most common strains In Europe [41], the US [13,42,42], and Australia [43] during the last decade. ST2 is commonly characterized in the literature as an endemic strain in the U.S. that has not been associated with hospital outbreaks [44]. However, it is also frequently classified as healthcare associated: the most recent data from the Centers for Disease Control and Prevention Emerging Infections Program reports between 41% and 52% of RT014 were considered healthcare-associated infections between 2012 and 2017 [13]. Despite this, evidence of transmission of RT014/ST2 within the hospital is sparse, as demonstrated by this study and others [6,13]. One explanation for this discordance between genomic evidence of recent transmission and healthcare-associated characterization via surveillance definitions is that ST2 is frequently acquired in the community, imported into the hospital, and subsequently progresses to infection after hospitalization. If this is the case, antimicrobial stewardship interventions may be particularly effective for preventing infections due to this common strain [7]. Environmental studies that have reported recovery of RT014 isolates in agriculture [45,46], wastewater [47], and parks and homes [39] are also consistent with community circulation of RT014. Overall, this finding highlights the imperfect nature of relying on infection onset as a proxy for acquisition. With the advent of more widespread pathogen whole genome sequencing, genomic evidence of healthcare transmission could be used as an alternative and more accurate metric than infection onset for measuring within-hospital transmission of *C. difficile*.

We also observed a notable difference in concordance between genomic linkages (isolate related within small SNV distance thresholds) and epidemiologic linkages (isolates collected from the same site within temporally proximate time periods) among ST1 and ST2 isolates. Specifically, ST2 isolates were more likely to have close genomic neighbors across disparate geographic sites and long time periods. Consistent with this, a pan-European surveillance study reported that the average most closely related strain to any given RT014 isolate was collected from hundreds of miles away [13]. The mechanisms behind this finding are not clear, but are consistent with a reliance on non-healthcare routes of spread. Practically speaking, this finding highlights the risks of broadly applying SNV thresholds to infer recent transmission, even to isolates of the same species. In particular, it emphasizes the importance of considering background genomic diversity and incorporating geographically and temporally diverse strains when interpreting genomic linkages. Without this context, one might mistakenly attribute a linkage to transmission when it in fact reflects broader genomic diversity patterns in a particular lineage. The importance of genomic context has been noted since the early days of bacterial genomic epidemiology [48], but in most cases, sequencing is still not widespread enough to provide such context. As we continue to consider a future with routine genomic surveillance in hospital settings to identify outbreaks [49], it is crucial that assessment of genomic context remain part of the evidence required for inferring transmission from genomic data.

*C. difficile*’s spore-forming lifestyle may contribute to some of the results reported here. It has been posited that spore formation likely drags down average estimate evolutionary rates of bacteria [50]. Extending from that, if isolates belonging to particular lineages spend more time in spore form than others, that lineage could be expected to have a lower average evolutionary rate, and thus, less nucleotide differences accumulated over time. We speculate that the ST2 and FQS ST1 lineages may have spent, on average, more time in spore-form than the epidemic and more recently emerged FQR ST1 lineages resulting in more closely related isolates across larger amounts of time and space. Ecological niches may influence this: more selective pressures and a higher density of susceptible hosts in healthcare settings could facilitate more time in the vegetative state, whereas strains that circulate primarily in the community may be more likely to stay dormant for longer periods of time. Results from our Bayesian timed phylogenomic analyses were consistent with this framework in two ways: 1) high evolutionary rate heterogeneity in both ST1 and ST2 isolates may reflect the effects of spore formation, with isolates emerging for a long-dormant spore being found on the tips of phylogenetic branches with a slow estimated evolutionary rate and 2) less evidence of temporal signal and slightly lower estimated evolutionary rates for FQS-ST1 isolates and ST2 isolates compared to FQR-ST1 isolates may reflect more time spent in spore-form. Whatever the biological and epidemiological underpinnings of the patterns we observed, this work highlights the challenges inherent to applying molecular clock-based methods to studying the epidemiology and evolution of a variably and relatively slowly evolving pathogen like *C. difficile*.

Our findings should be interpreted in the context of multiple limitations. First, the retrospective nature of the study resulted in some differences in sample collection between the three study sites: UM and TMC selected based off of PCR Ribotypes, which we then filtered down to only ST1 and ST2 via *in silico* MLST, while MSKCC originally selected isolates based off of ST as MLST is routine at that center. However, all comparisons were made between ST1 and ST2 isolates and these differences were consistent within the ST1 and ST2 isolates at each site, so we would not expect them to significantly alter the results reported here. Second, limited epidemiologic metadata was available for analysis: only study site and collection date. Despite this, the interesting patterns we observed between genomic linkages and epidemiologic linkages emphasizes the value of integrating genomic data with even limited epidemiologic metadata. Finally, the evolutionary rate estimates presented here are subject to uncertainty, particularly given the observed instances of violated model assumptions and relatively limited temporal signal in the data. However, the overall trends remained stable with varying models, alleviating concerns that our findings are artifacts of model misspecification. This study also has several notable strengths, including the collection of isolates from three distinct geographic sites in the U.S., the application of whole genome sequencing for high-resolution typing and phylogenetic analyses, and the incorporation of global isolates for increased context and power in our timed phylogenomic analyses.

## Conclusions

Examination of the genomic epidemiology of *C. difficile* ST1 and ST2 across three geographically distinct U.S. medical centers revealed divergent epidemiologic and evolutionary patterns between these two common strains. Specifically, we observed more evidence of geographic clustering, recent healthcare transmission, and a slightly more rapid average evolutionary rate among FQR ST1 isolates compared to ST2 and FQS ST1 isolates. One implication of these findings is that an understanding of local molecular epidemiology may facilitate the development of effective interventions targeted at reducing the burden of CDI. These findings also highlight how methodological considerations—including incorporating genomic context when inferring transmission from genomic linkages and considering the potential effect of spore formation on the connection between genomic differences and epidemiology—need to be accounted for when applying genomic epidemiology methods for studying *C. difficile* transmission.

## Data Availability

All whole genome sequence data was uploaded to the National Center for Biotechnology Information (NCBI) Sequence Read Archive (SRA) under Bioproject accessions PRJNA595724, PRJNA561087, and PRJNA594943. Metadata that comply with patient privacy rules are included in the Supplementary Materials.

## ACKNOWLEDGEMENTS

We thank Ali Pirani for his contributions to the bioinformatics data analyses.

