## Supplementary Materials for "A multisite genomic epidemiology study of *Clostridioides difficile* infections in the U.S. supports differential roles of healthcare versus community spread for two common strains"

**Supplementary Figure 1:** Dot plot of newly sequenced ST1 and ST2 isolates by collection date and study site. While collection timing varied by study site, timing was similar between ST1 and ST2 isolates. MSKCC = Memorial Sloan Kettering Cancer Center, TMC = Texas Medical Center Hospital, UM = Michigan Medicine.

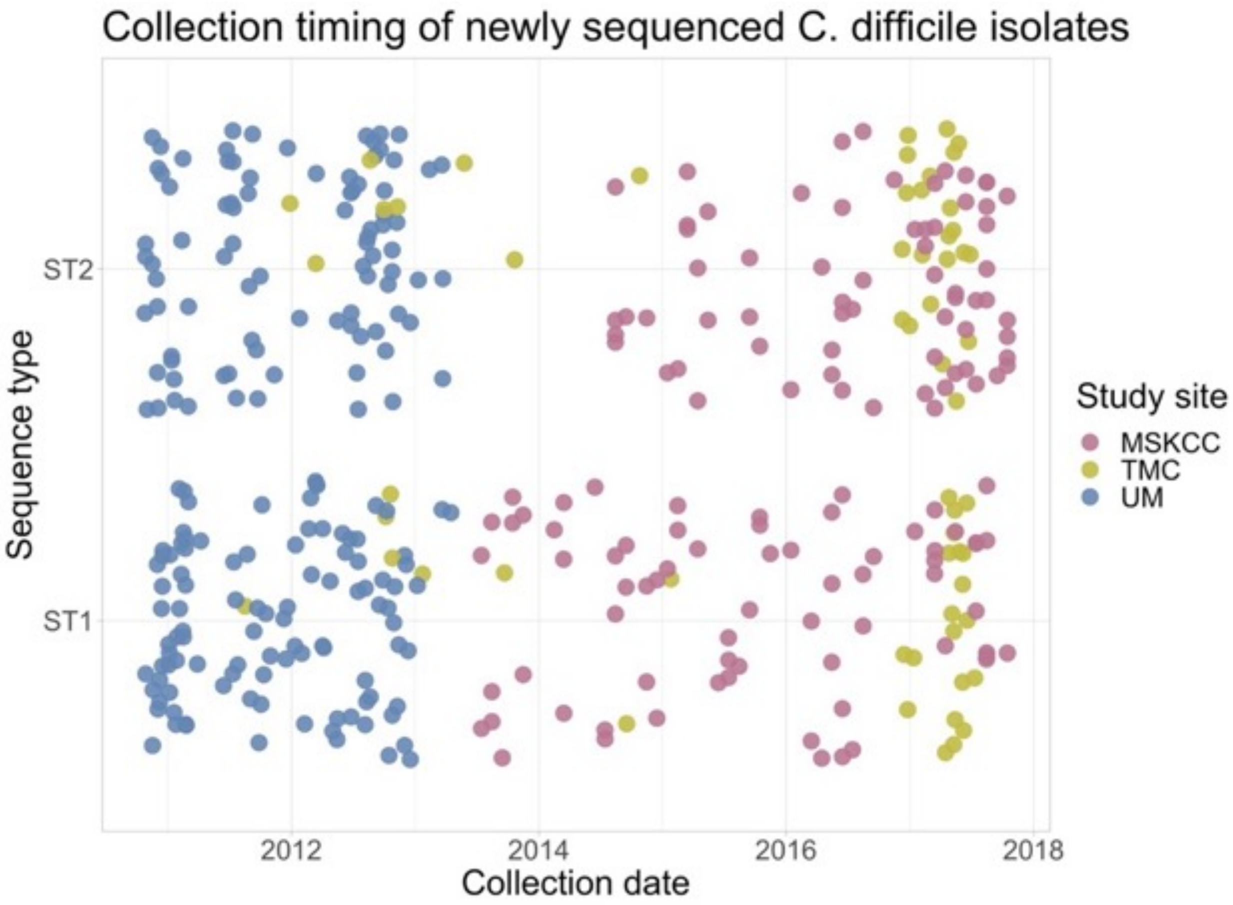

**Supplementary Figure 2:** Results of clustering permutation tests for clustering of study site on phylogenies composed of newly sequenced ST1 and ST2 isolates. Red diamond indicates metric of clustering on real data, while the distribution represents the range of clustering observed in 1,000 datasets with randomly permuted study sites. The further outside the distribution the real estimate is, the stronger the evidence that the observed clustering is more than would be observed by chance.

# ST1

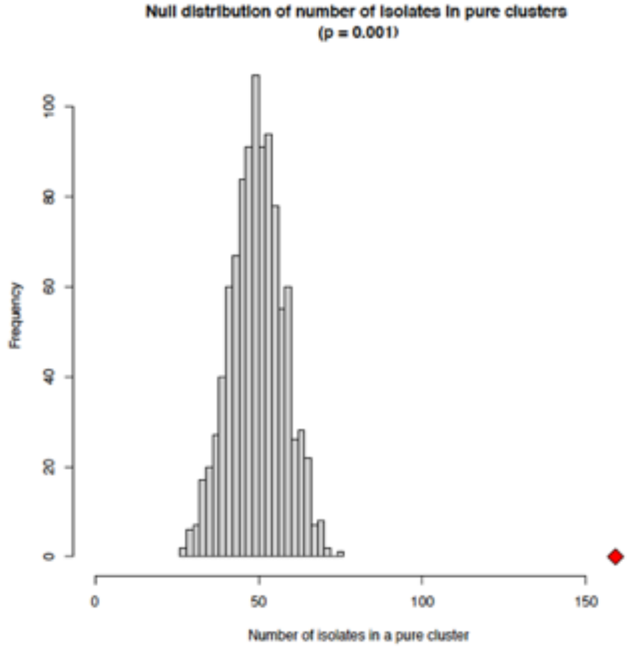


# ST2

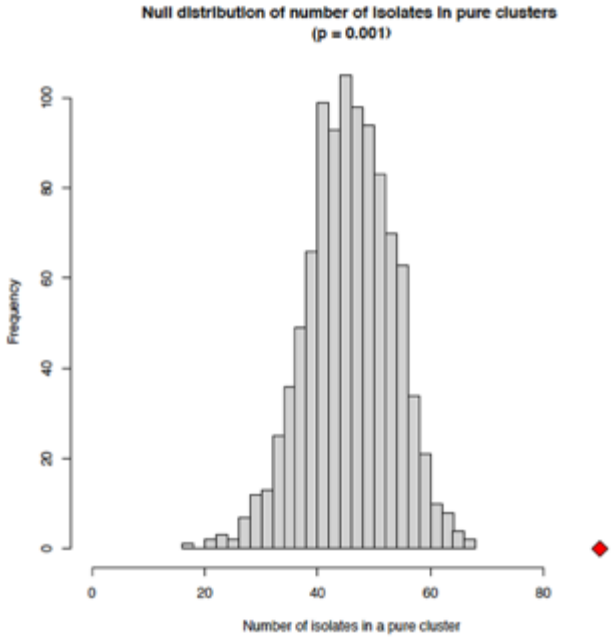

**Supplementary Figure 3:** Regression of sampling date on root-to-tip distance for ST1, ST2, and FQR ST1 and FQS ST1 only datasets. While ST1, ST2, and FQR ST1 have a positive correlation, indicating some temporal signal, FQS ST1 has a negative correlation, indicating a lack of temporal signal.

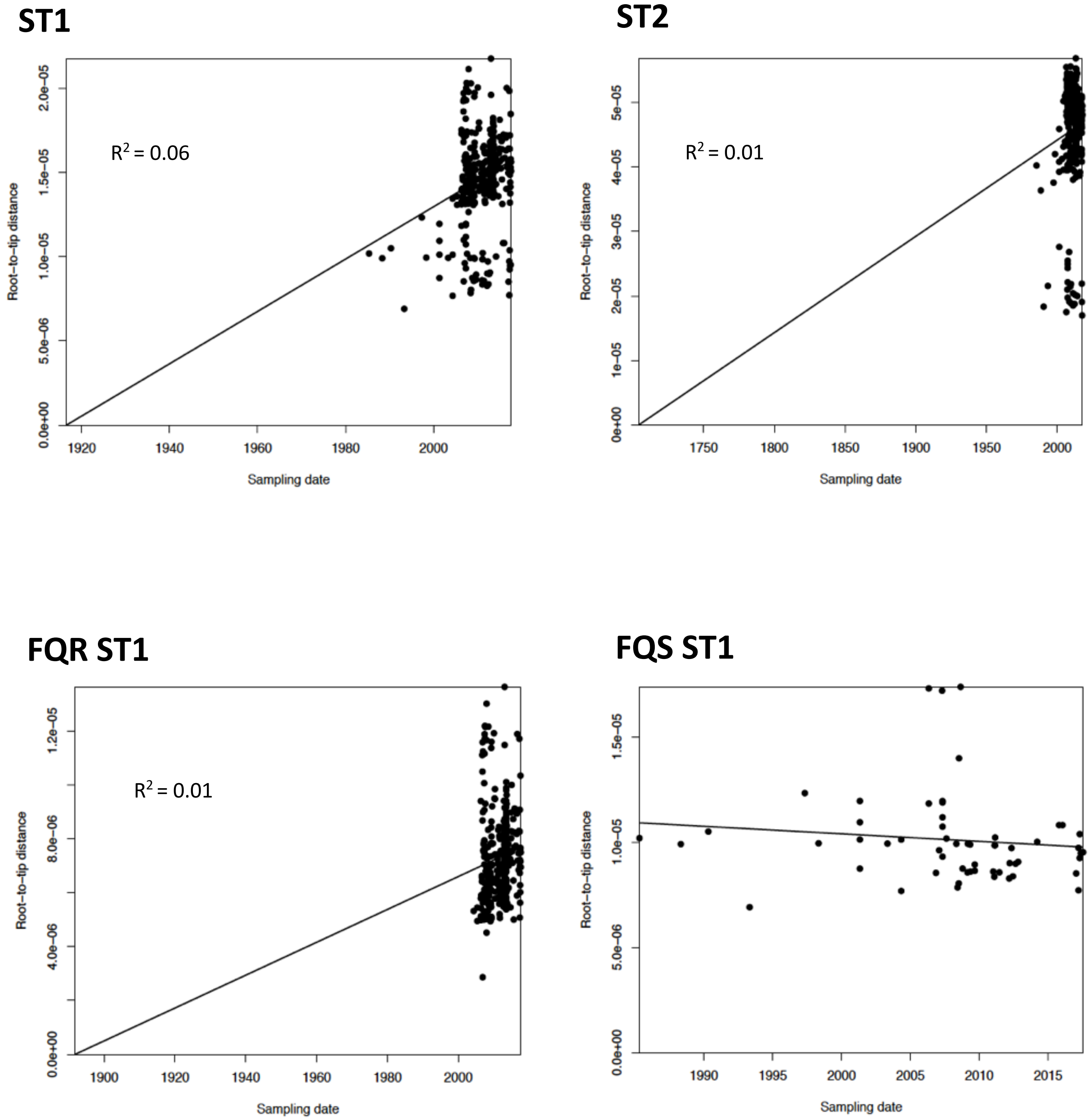

**Supplementary Figure 4:** Results of date randomization tests for each dataset; point furthest to the left is the evolutionary rate estimate for the real data, the rest of the points are evolutionary rate estimates from datasets with randomly permuted dates. Bars represent 95% highest posterior density intervals as estimated by BEAST.

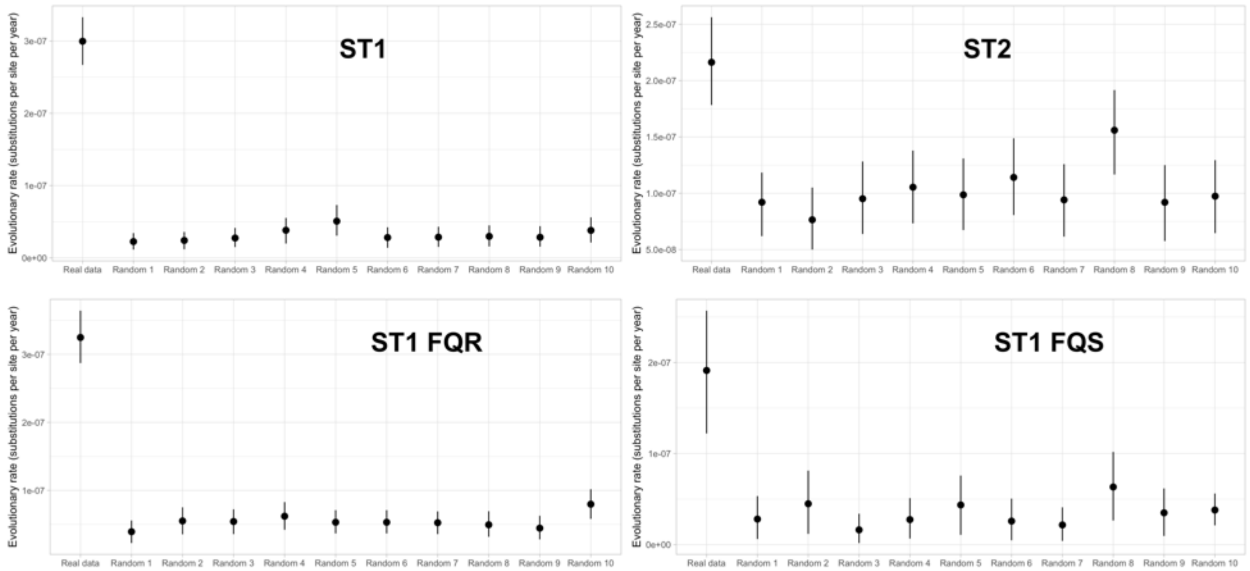

### Supplementary Results

#### *BEAST model selection*

Our model selection process for BEAST considered the bias-variance tradeoff of allowing for more complex molecular clock and demographic model priors given limited temporal signal in the data. When evaluating appropriate molecular clock models, the four examined datasets (all ST1, FQS ST1 only, FQR ST1 only, ST2) displayed varying degrees of evidence of evolutionary rate heterogeneity, violating the strict molecular clock assumption, with FQS ST1 isolates displaying the most evolutionary rate heterogeneity (Supplementary Figure 5A). This observation supported the application of uncorrelated relaxed lognormal clock models for every dataset, which was successfully implemented.

The evaluation of appropriate demographic priors was more complex. All four datasets demonstrated some evidence of a non-constant population size, violating the constant population demographic model, with FQR ST1 isolates violating this assumption to the greatest extent (Supplementary Figure 5B). As a result, we attempted to run non-parametric Gaussian Markov Random Field (GMRF) skyride demographic priors for all datasets. However, our data demonstrated limited ability to accommodate more complex and flexible demographic model prior assumptions. Specifically, we were either not able to successfully initialize the model runs (ST2 and All ST1 datasets) or the runs initialized but produced suspicious results (FQR-ST1 and FQS-ST1 datasets) in the form of unusually recent and precise time to most recent common ancestor estimates. These suspicious results raised concerns that the GMRF smoothing prior was overly influential and biasing the results, particularly given the limited temporal signal in the data [1]. Thus, we ultimately applied a constant population size demographic prior for all of the evolutionary rate estimates. However, although not selected for the main analyses, the evolutionary rate estimates from the executed GMRF skyride models displayed the same general trends as the main models, with FQR ST1 displaying evidence of a higher evolutionary rate than FQS ST1 and ST2 datasets (Supplementary Figure 6).

**Supplementary Figure 5:** (A) Density curves of coefficient of variation for all four datasets when applying an uncorrelated lognormal molecular clock and a constant population size. A higher correlation coefficient indicates greater violation of strict molecular clock assumption (B) Density curves of the exponential growth rate coefficient for all four datasets when applying an uncorrelated lognormal molecular clock and an exponential population size. A higher exponential growth rate indicates greater violation of the constant population size demographic prior.

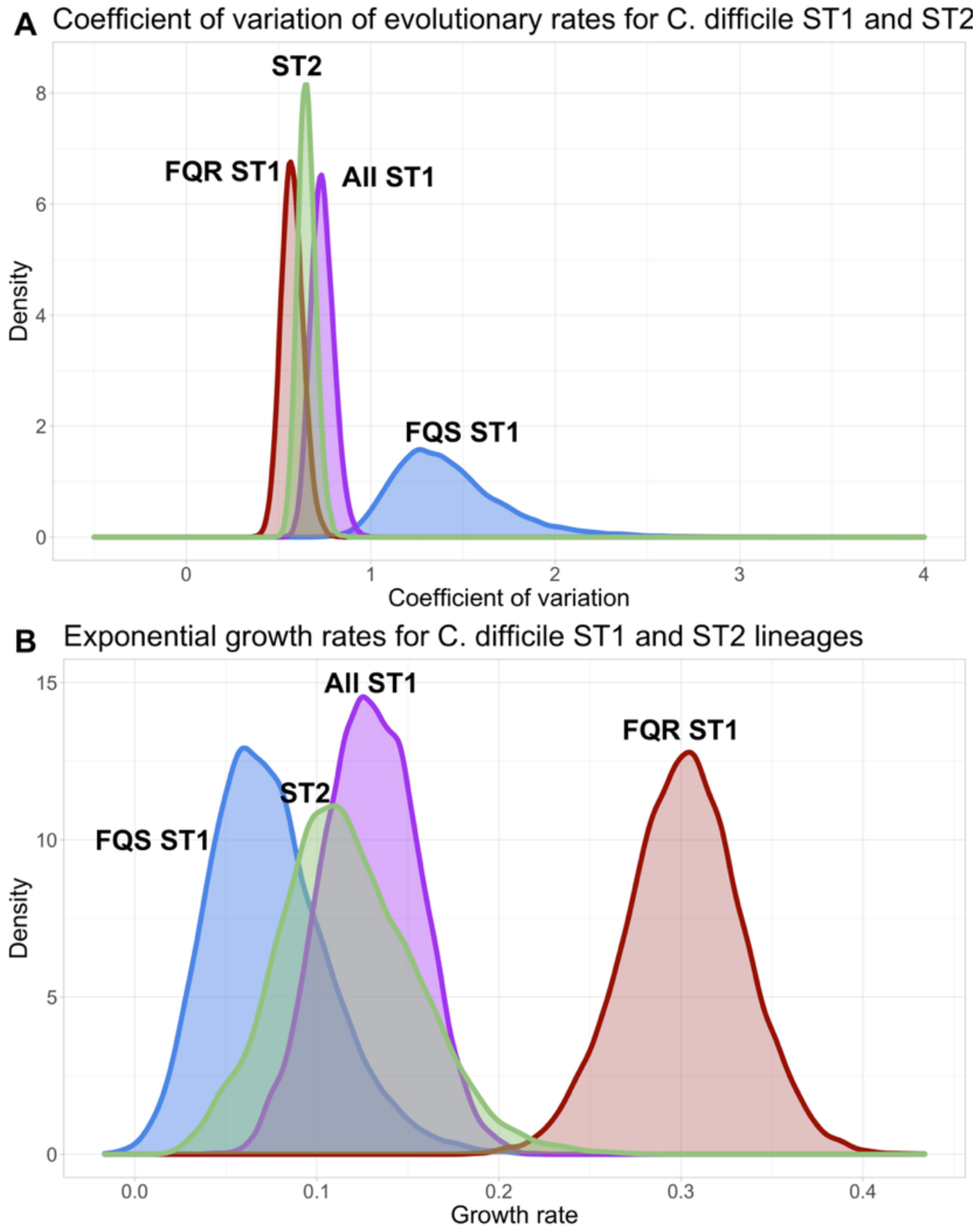

**Supplementary Figure 6:** Evolutionary rate estimates for ST1 and ST2 *C. difficile* lineages when applying non-parametric Gaussian Markov random field (GMRF) skyride demographic priors to the FQR ST1 and FQS ST1 datasets (reflected with dotted lines). The ST2 dataset was run with a constant demographic prior model for ST2 because the GMRF model would not initialize. The same overall trends observed in the main analysis (when all models used a constant population demographic prior) persist.

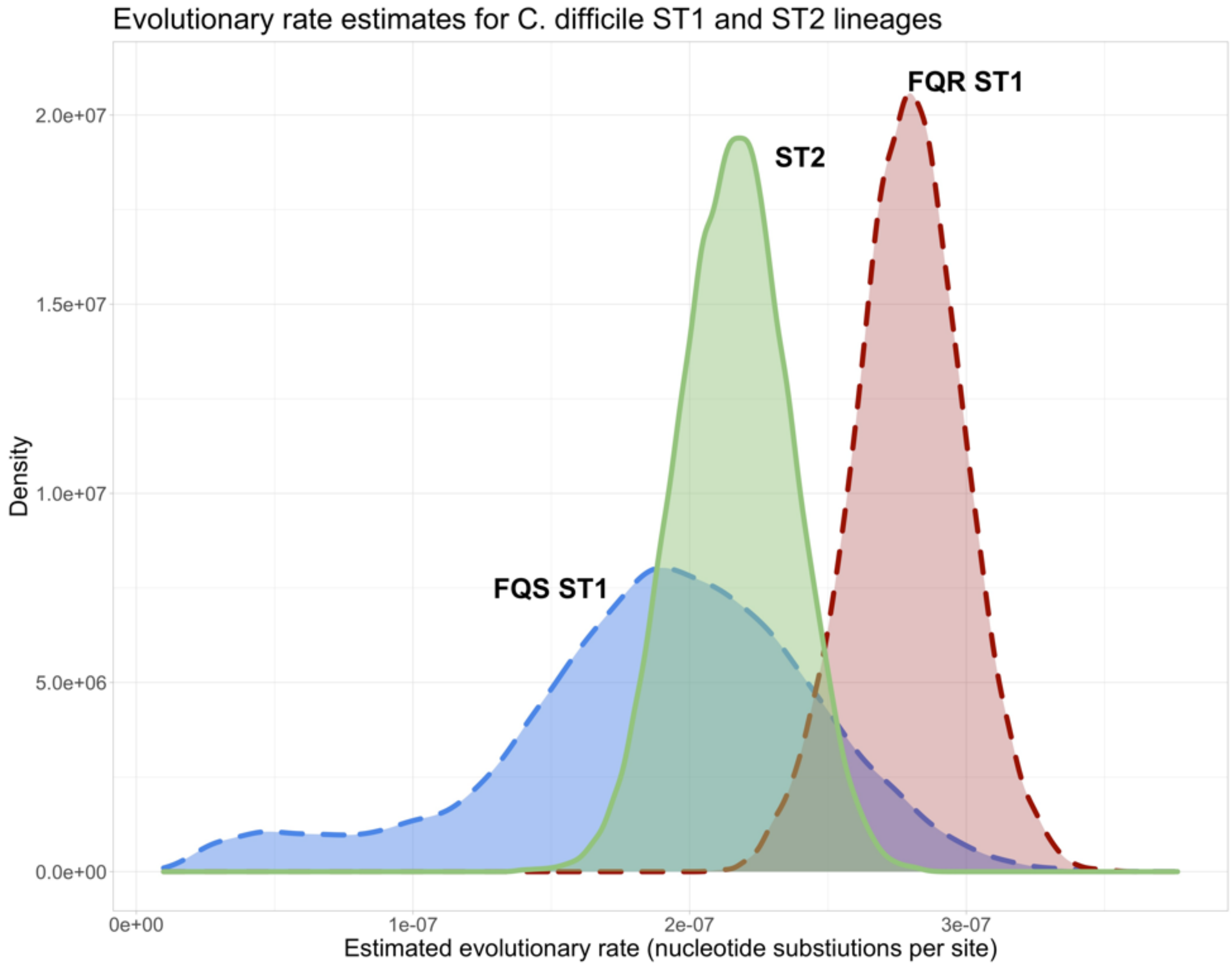
